# Assessment of COVID-19 risk and prevention effectiveness among spectators of mass gathering events

**DOI:** 10.1101/2021.07.05.21259882

**Authors:** Tetsuo Yasutaka, Michio Murakami, Yuichi Iwasaki, Wataru Naito, Masaki Onishi, Tsukasa Fujita, Seiya Imoto

## Abstract

There is a need to evaluate and minimise the risk of novel coronavirus infections at mass gathering events, such as sports. In particular, to consider how to hold mass gathering events, it is important to clarify how the local infection prevalence, the number of spectators, the capacity proportion, and the implementation of preventions affect the infection risk. In this study, we used an environmental exposure model to analyse the relationship between infection risk and infection prevalence, the number of spectators, and the capacity proportion at mass gathering events in football and baseball games. In addition to assessing risk reduction through the implementation of various preventive measures, we assessed how face-mask-wearing proportion affects infection risk. Furthermore, the model was applied to estimate the number of infectors who entered the stadium and the number of newly infected individuals, and to compare them with actual reported cases. The model analysis revealed an 86%–95% reduction in the infection risk due to the implementation of face-mask wearing and hand washing. Among the individual measures, face-mask wearing was particularly effective, and the infection risk increased as the face-mask-wearing proportion decreased. A linear relationship was observed between infection risk at mass gathering events and the infection prevalence. Furthermore, the number of newly infected individuals was also dependent on the number of spectators and the capacity proportion independent of the infection prevalence, confirming the importance of considering spectator capacity in infection risk management. These results highlight that it is beneficial for organisers to ensure prevention compliance and to mitigate or limit the number of spectators according to the prevalence of local infection. Both the estimated and reported numbers of newly infected individuals after the events were small, below 10 per 3–4 million spectators, despite a small gap between these numbers.

## 1. Introduction

Since the global pandemic of corona virus disease 2019 (COVID-19), various measures have been taken, ranging from those related to individual behavioural changes, such as physical distance and face-mask wearing, to community-wide measures, such as lockdowns (*1, 2*). Mass gathering events were considered to be a factor in the spread of the disease (*3-5*), forcing cancelation, postponement, or behind-closed-doors games of various sporting events. It was also pointed out that restrictions on mass gathering events might contribute to the suppression of infection, because stricter restrictions on the number of spectators at mass gathering events were likely to result in a lower effective reproduction number of the virus for the following two weeks (*6*). Since then, along with the implementation of infection preventive measures, a variety of sporting events have been held with gradually fluctuating capacity proportions (i.e., attendance rates). For example, the Australian Open tennis tournament in February 2021 was held with about 50% spectators of the stadium’s capacity for the last four days of the events. During the 2020 season (February 2020–January 2021) in Japan, the total number of spectators was 4,823,578 in professional baseball games (*7*) and 3,615,066 in professional football games (*8*), but no secondary infection was observed among the spectators. The United Kingdom, Spain, and other countries have been conducting empirical studies on mass gathering events since December 2020 (*9, 10*).

By contrary, the risk of COVID-19 infection among spectators at mass gathering events remains unclear. Although there have been reports on estimates of the probability of infected individuals entering among spectators (*11, 12*) and tools for risk assessment and mitigation of mass gathering events (*13*), limited studies are available on infection risk caused by mass gathering events. Murakami et al. recently developed an environmental exposure model for infection risk at mass gathering events and applied it to an assessment of infection risk among spectators and the reduction effect of preventions at the opening ceremony of the Tokyo 2020 Olympic Games (*14*). However, how the infection prevalence among entering spectators, the number of spectators, the capacity proportion, and the implementation of preventive measures affect the infection risk has not been unravelled. Clarifying the relationship between infection risk and these factors is important for future discussions on the regulation and mitigation of the number of spectators and infection prevention compliance at mass gathering events.

In this study, we applied the environmental exposure model to simulate actual baseball and football game conditions for the assessment of infection risk among spectators by varying the infection prevalence among spectators, the number of spectators, and the capacity proportion. We also evaluated the reduction effect of preventive measures by comparing the infection risk with and without preventions. Furthermore, we clarified the relationship between infection risk and the proportion of face-mask wearing. Lastly, we applied the model and the local infection prevalence to estimate the total number of infectors who entered the stadium and the total expected number of -newly infected individuals in the 2020 season, and to compare them with the actual reported cases.

## 2. Methods

### 2.1 Model

In this study, we used the environmental exposure model (*14*) with small modifications. Briefly, the model simulates the infection risk from four pathways: direct exposure through droplet sprays, direct exposure from inhalation of inspirable particles, hand-to-face contact exposure contaminating mucous surfaces, and inhalation of respirable particles via air (*15*). It allows for the effectiveness of preventions in reducing the risk of infection by assessing the exposure by pathways. Given the probability of asymptomatic infectors of COVID-19 among spectators entering a stadium (i.e., infection prevalence, *P*_0_), we calculated the viruses released by infectors during coughing, talking, and sneezing, as well as their environmental behaviour, inactivation, and surface transfer. Parameters included frequency of coughing, talking, and sneezing, saliva volume (*16-19*), virus concentration in saliva, viral viability ratio (*20-22*), environmental inactivation rate (*23*), air exchange rate, breath volume rate (*24*), distance between the persons, frequency of contacts to the facial surface (*25*), surface transfer coefficient (*15*), and dose-response model (*26*). There were five categories of spectators: (1) those who accompanied the infectors, (2) those who sat in front of the infectors in the stand, (3) those who used the restroom after the infectors used it, (4) those who ordered after the infectors ordered at the concessions, and (5) others. We assumed that all non-infectors were susceptible to the viruses. The stadium was divided into two locations: the stands and the rest of the stadium (i.e., concourses, concessions, and restrooms).

There were four modifications to the previous model (*14*). First, in the previous model, we assumed that people spent 15 minutes to enter the stadium, 15 minutes to use the restroom, 15 minutes to order at the concessions, 4 hours in the stands, and 15 minutes to leave the stadium. However, in this study, since we simulated a football or baseball game, we assumed that people spent 1 hour in the football or baseball stands before the game, 2 hours in the football game, and 3 hours and 10 minutes in the baseball game (i.e., the total time spent in the stands was changed to 3 hours for the football game and 4 hours and 10 minutes for the baseball game). Second, the number of accompanies with the infector was set at a 50% probability of two, a 35% probability of one, and a 15% probability of zero (i.e., the infector alone) based on realistic numbers of accompaniers (*27, 28*). The probability of an infector talking toward an accompanier in the stands was set at 0.3 before and 0.2 during the game for the no-prevention scenarios, and at 0.15 before and 0.1 during the game under the condition of the presence of preventive measures. The probabilities of coughing, sneezing, and talking outside the stands were the same as those used in the previous model. Third, the following seven preventive measures were assumed in the previous model: (a) physical distance during entry and exit, (b) decontamination of environmental surfaces, (c) stadium air ventilation, (d) partitioning between spectator seats, (e) face masks use, (f) hand washing, and (g) wearing hats or other headwear. However, we assumed that the partitioning of the spectators in the stands was not used, and face masks were to be worn even in the stands instead. In Condition A (see below for details), these six measures were considered, while in Condition B, only face masks and hand washing were considered. Studies have suggested that wearing a face mask eliminates 95% of the virus attached to large particles (with an aerodynamic diameter of > 10 µm) emitted by infectors (*29, 30*) and that it also reduces the frequency of facial mucosal membrane touches by 67% (*25*). Fourth, the parameters of *P*_0_, stadium capacity (i.e., the maximum number of spectators), capacity proportion (i.e., the ratio of the number of spectators to the stadium capacity), and face-mask-wearing proportion were varied as experimental conditions in this study. The distance between the infector and the accompanier in the stands was set to a value corresponding to the capacity proportion. Specifically, the distance was 0.5 m at a capacity proportion of 100%, 1 m at 50%, and 1.5 m at 33% or lower. When the capacity proportion was between 50% and 100%, it was either 0.5 m or 1 m, depending on the capacity proportion; when the capacity proportion was between 33% and 50%, it was either 1 m or 1.5 m. Similarly, the distance between the infector and the person sitting in front of the infector in the stands was also set to a value, depending on the capacity proportion. When the capacity proportion was 100%, the distance was set to 0.5 m. The distance was set to 1 m at a capacity proportion of less than 50%. When the capacity proportion was between 50% and 100%, the value was either 0.5 m or 1 m, depending on the capacity proportion.

### 2.2. Conditions

In this study, we first evaluated the effectiveness of individual or combined preventive measures, described as Condition A. Under the conditions of a football game, *P*_0_ = 10^−3^, stadium capacity = 40,000 persons, and capacity proportion = 50% (20,000 spectators), eight conditions were analysed: no preventions, six individual measures, and all combined measures. Assuming that infectors remain infective for an average of 9.3 days (*31*), that the ratio of asymptomatic infected individuals to the number of all infected individuals is 46% (*32*), and that symptomatic individuals experience 2.3 days of infectivity before symptom onset (*31*), *P*_0_ = 10^−3^ represented 1,800 newly infected individuals, including both asymptomatic and symptomatic cases per day (12,700 per week) in the population of 10 million people. The risk of infection with all the six preventive measures implemented was also analysed by varying the face-mask-wearing proportion. The face-mask-wearing proportion was 0%, 10%, 20%, 30%, 40%, 50%, 60%, 70%, 80%, 90%, and 100% (the condition of 100% face-mask-wearing proportion was the same as that with all measures described above). Infectors were assumed to be wearing face masks according to these probabilities. In this case, all measures other than face masks were considered to be implemented. It should be noted that the average percentage of spectators wearing face masks in football games has been observed to be over 90% (*33*).

Next, as Condition B, we examined the infection risk depending on *P*_0_, the stadium capacity, the capacity proportion, and the presence or absence of preventions. There were six conditions for *P*_0_ (10^−6^, 10^−5^, 2×10^−5^, 10^−4^, 2×10^−4^, 10^−3^), five conditions for the stadium capacity (5,000, 10,000, 20,000, 40,000, and 80,000 persons), four conditions for the capacity proportion (25%, 50%, 75%, and 100%), two conditions for preventions (presence and absence of measures), and two sports (football and baseball games). As preventive measures, a combination of face mask wearing and hand washing was considered. We analysed a total of 480 conditions (6 × 5 × 4 × 2 × 2). Monte Carlo simulations were performed 10,000 times per condition to calculate the expected infection risk per spectator other than infectors and the expected number of newly infected individuals.

A log-normal generalised linear model (GLM) (i.e., a GLM with a normal distribution and log-link function) was used to predict the expected number of newly infected individuals at Condition B with preventive measures (*Num*_*inf*_) using three explanatory variables: *P*_0_, the number of speculators (*Num_speculators*), and the capacity proportion (*Percent*). Log_10_-transformed values of *P*_0_ and *Num_speculators* were used in this modelling. The model formula was as follows:

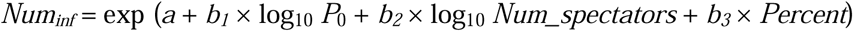

where *a*_*1*_ is the intercept, *b*_*1–3*_ are the partial regression coefficients for the explanatory variables.

Using maximum likelihood estimation, we developed two regression models separately for football and baseball games. To compare the relative importance among the explanatory variables, we re-estimated the partial regression coefficients by using those variables standardised. We did not calculate so-called “standardised partial regression coefficients” because the standardization of the objective variable was not applicable in the GLM estimation. The GLM estimation was performed by using the “glm” function in the “stats” package of R version 4.0.5 (*34*).

We then estimated the total expected number of newly infected individuals during 2020 seasons after August 2020 (J. League professional football: 878 games until January 4, 2021, with a total of 2,935,947 spectators; professional baseball: 512 games until November 25, 2020, with a total of 4,439,258 spectators) by applying *P*_0_, the number of spectators, and the capacity proportion at each game. Based on the cumulative number of confirmed cases per week, including the game day in the prefecture where the game was held (*35*), *P*_0_ was calculated by assuming that *P*_0_ = 10^−3^ corresponded to 1,800 new infection cases/day (12,700 infection cases/week) in the population of 10 million people. Since the number of cases confirmed by testing may be less than the actual number of newly infected individuals, the calculated *P*_0_ was likely to be smaller than the actual value.

Monte Carlo simulation and multiple regression analysis were performed using R (*34*).

### 2.3. Cautions for interpretations and uncertainties

The interpretation of the results of this study requires some cautions. First, it was assumed that there were no antibody carriers. If half of the audience had antibodies, the risk of infection would be halved. Therefore, an increase in the number of antibody carriers would contribute to a reduction in the risk of infection in the stadium. Second, because we aimed to estimate the risk of infection in football and baseball games during the 2020 season, the increased risk of infection due to mutant strains was not taken into account. One possible way to estimate the risk associated with mutant strains is to change the virus concentration in saliva. When the virus concentration in saliva was changed to 10 and 100 times, the risk of infection was 3.1 and 12 times, respectively, without preventions, and 6.9 and 24 times, respectively, with preventions (Condition B) under the conditions of *P*_0_ = 10^−3^, the stadium capacity = 40,000, and the capacity proportion = 50% (Figure S1). Third, this simulation results showed the arithmetic mean of the Monte Carlo simulations in the set scenarios. Scenarios that were not set up (e.g., eating and drinking unmasked in a concourse or talking in a group) were out of the risk assessment. Similarly, the study identified the risk of infection inside the stadium, and did not cover the risk of infection outside the stadium, which may occur due to movement or concentration of people.

This study had some uncertainties, the details of which has been described in the previous study (*14*). In brief, the parameters used in this study are those that are valid for current knowledge, but there are certain uncertainties. Specifically, since the viral viability ratio of SARS-CoV-2 in human saliva was unknown, we used the value for a ferret (*21*). Similarly, we used a dose-response model for SARS-CoV developed using a mouse (*26*). Furthermore, we did not take into account the difference in saliva volume depending on the loudness of voice, the virus inactivation in fingers and hair, or the removal of viruses attached to small particles by face masks

## 3. Results

### 3.1. Preventive measure effectiveness

In condition A at a football game (*P*_0_ = 10^−3^, stadium capacity = 40,000, capacity proportion = 50%), the estimated infection risk was 7.4×10^−4^ for no preventions, 4.4×10^−4^ for physical distance, 5.6×10^−4^ for decontamination of environmental surfaces, 6.5×10^−4^ for ventilation, 5.5×10^−5^ for face masks, 6.5×10^−4^ for hand washing, 6.5×10^−4^ for headwear, and 2.2×10^−5^ for all six preventions (Figure 1a). As a single measure, the risk-reduction effect of wearing a mask was as high as 93%. The risk reduction effect was 97% when all six preventive measures were implemented.

The infection risk increased as the face-mask-wearing proportion decreased (Figure 1b). The infection risk was 2.5×10^−4^ when the face-mask-wearing proportion was 0% (i.e., other measures were implemented). The infection risk at a face-mask-wearing proportion of 90%, which is close to the value in actual games (*33*), was 4.9×10^−5^, which was 2.2 times higher than that at a face-mask-wearing proportion of 100%.

We then evaluated the risk reduction effect of the preventive measures at different *P*_0_, stadium capacities, and capacity proportions under condition B (Tables 1 and S1). Here, we show the reduction ratio of infection risk by implementing the measures of face-masking and hand-washing against the infection risk without measures at the same condition regarding *P*_0_, stadium capacity, and capacity proportion. The reduction ratios were in the range of 86%–95% for the football game condition and 90%–95% for the baseball game condition, although there were some differences in *P*_0_, stadium capacity, and capacity proportion.

**Figure 1.**
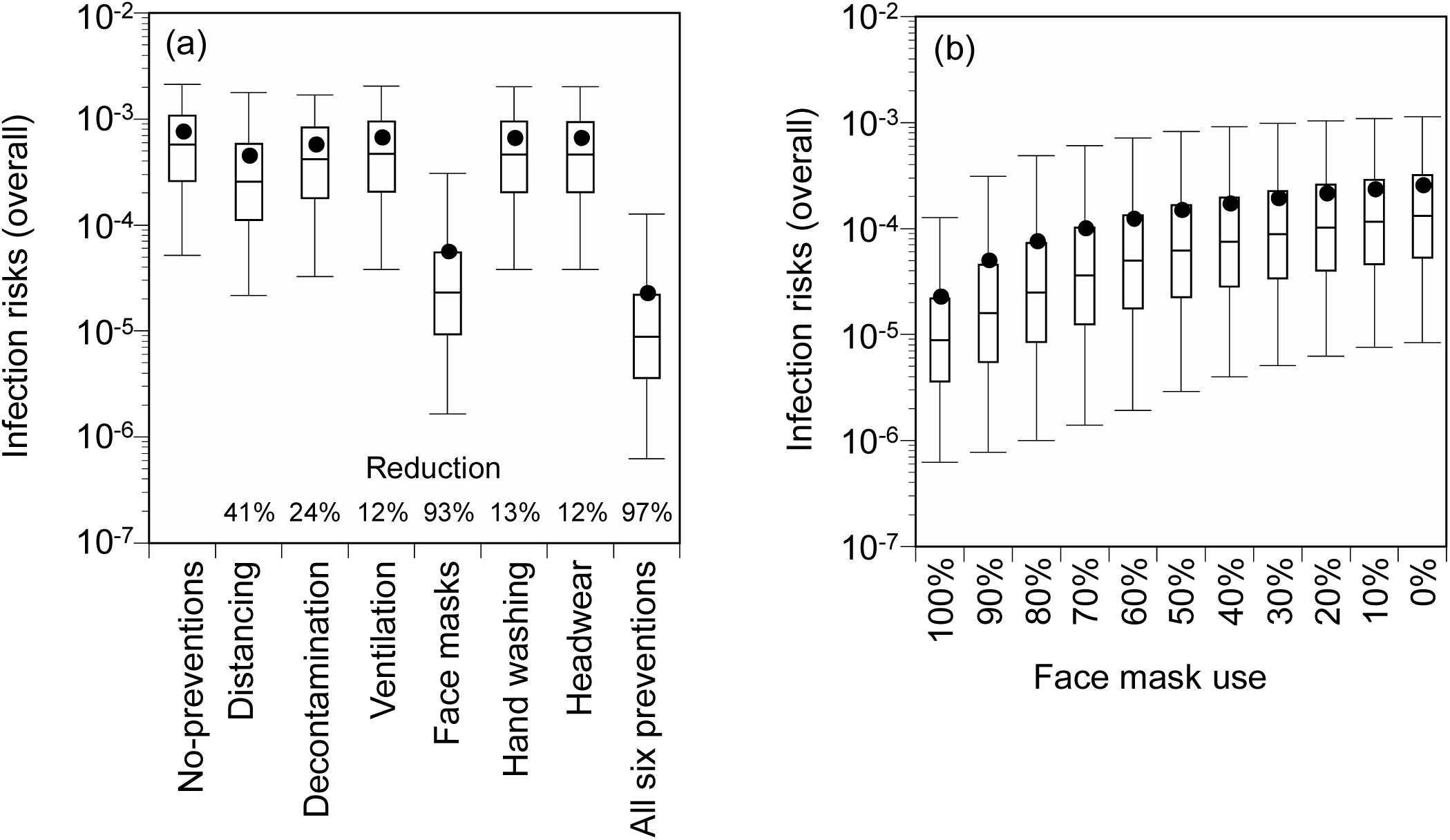
Comparison of infection risk among different preventive measures (Condition A). (a) Comparison of infection risk by each individual preventive measure and all six measures, (b) relationship between face-mask-wearing proportion and infection risk. Box-and-whisker plots represent the 2.5, 25, 50, 75, and 97.5 percentiles. Closed circles represent the arithmetic mean of the simulation.

**Table 1.** Reduction ratio of infection risks due to preventions at different *P*_0_, stadium capacity, and capacity proportion. Condition B, football condition. *P*_0_: a crude probability of a spectator being an infector.

| Stadium capacity | Capacity proportion | Number of spectators | $P_0$ | | | | | |
| --- | --- | --- | --- | --- | --- | --- | --- | --- |
| | | | $10^{-6}$ | $10^{-5}$ | $2 \times 10^{-5}$ | $10^{-4}$ | $2 \times 10^{-4}$ | $10^{-3}$ |
| 80000 | 100% | 80000 | 90% | 91% | 91% | 91% | 91% | 91% |
|  | 75% | 60000 | 92% | 92% | 92% | 92% | 92% | 92% |
|  | 50% | 40000 | 93% | 93% | 93% | 93% | 93% | 93% |
|  | 25% | 20000 | 93% | 93% | 93% | 93% | 93% | 93% |
| 40000 | 100% | 40000 | 90% | 91% | 91% | 91% | 91% | 91% |
|  | 75% | 30000 | 92% | 92% | 92% | 92% | 92% | 92% |
|  | 50% | 20000 | 93% | 93% | 93% | 93% | 93% | 93% |
|  | 25% | 10000 | 92% | 93% | 93% | 93% | 93% | 93% |
| 20000 | 100% | 20000 | 89% | 91% | 91% | 91% | 91% | 91% |
|  | 75% | 15000 | 91% | 92% | 92% | 92% | 92% | 92% |
|  | 50% | 10000 | 91% | 93% | 93% | 93% | 93% | 93% |
|  | 25% | 5000 | 92% | 94% | 93% | 93% | 93% | 93% |
| 10000 | 100% | 10000 | 88% | 90% | 91% | 91% | 91% | 91% |
|  | 75% | 7500 | 90% | 92% | 92% | 92% | 92% | 92% |
|  | 50% | 5000 | 91% | 93% | 93% | 93% | 93% | 93% |
|  | 25% | 2500 | 91% | 93% | 94% | 93% | 93% | 93% |
| 5000 | 100% | 5000 | 86% | 90% | 90% | 91% | 91% | 91% |
|  | 75% | 3750 | 88% | 92% | 92% | 92% | 92% | 92% |
|  | 50% | 2500 | 89% | 93% | 93% | 93% | 93% | 93% |
|  | 25% | 1250 | 95% | 93% | 93% | 93% | 93% | 93% |

### 3.2. Relationship between infection risk and *P*_0_, stadium capacity, and capacity proportion

We compared the infection risk under Condition B among different *P*_0_ (Figures 2 and S2): stadium capacity = 40,000, capacity proportion = 50%, and the presence of measures. Under a football game condition, the infection risk at *P*_0_ = 10^−6^ was 4.8×10^−8^, while it was 5.3×10^−7^ at *P*_0_ = 10^−5^, 5.2×10^−6^ at *P*_0_ = 10^−4^, and 5.2×10^−5^ at *P*_0_ = 10^−3^. The infection risk increased by a factor of 1100 when *P*_0_ increased 1000 times. The similar results were obtained for the baseball game condition.

We then compared the expected number of newly infected individuals among different stadium capacities and capacity proportions at *P*_0_ = 10^−3^ and presence of measures (Figures 3 and S3). Under the stadium capacity = 80,000 persons and a football game condition, the expected number of newly infected individuals was 0.89 at the capacity proportion = 25% and increased to 7.2 by a factor of 8.0 at the capacity proportion = 100%. Similarly, the ratio of the expected number of newly infected individuals at the capacity proportion of 100% to 25% at the stadium capacity of 40,000, 20,000, 10,000, and 5,000 ranged from 7.9 to 8.0. The expected number of newly infected individuals at the stadium capacity = 80,000 persons and the capacity proportion = 50% (40,000 spectators) was 2.1, whereas that at the stadium capacity = 40,000, and the capacity proportion = 100% (40,000 spectators) was 3.6. Similarly, when comparing the expected number of newly infected individuals under the three conditions of the same number of spectators (20,000 persons), the condition with the stadium capacity of 80,000 persons and the capacity proportion of 25% showed the lowest at 0.89, followed by 1.0 under the stadium capacity of 40,000 persons and the capacity proportion of 50%, and 1.8 under the stadium capacity of 20,000 persons and the capacity proportion of 100%. Similar results were obtained for the same number of spectators with the different stadium capacities and the capacity proportions (i.e., 10,000 spectators: stadium capacity of the stadium capacity of 40,000 persons and the capacity proportion of 25%, 20,000 persons and 50%, and 10,000 persons and 100%; 5,000 spectators: stadium capacity of the stadium capacity of 20,000 persons and the capacity proportion of 25%, 10,000 persons and 50%, and 5,000 persons and 100%): the expected number of newly infected individuals slightly increased with an increase in the capacity proportion. Similar results were also confirmed in the baseball game condition.

Figure 4 shows the relative risk of infection with the preventive measures at different *P*_0_ and capacity proportions in comparison to the infection risk at *P*_0_ = 10^−3^, capacity proportion = 100%, and absence of measures (the stadium capacity = 80,000 persons; football game condition). There was a large reduction in the relative risk of infection due to the implementation of the preventive measures (e.g., 0.093 relative risk of infection with the presence of the measures at *P*_0_ =10^−3^ and capacity proportion = 100%). There was a large variation in the relative risk of infection due to different *P*_0_ (e.g., under conditions with presence of measures and capacity proportion = 100%: 0.0093 at *P*_0_ = 10^−4^; 0.00094 at *P*_0_ = 10^−5^; 0.000096 at *P*_0_ = 10^−6^) and a decrease in relative infection risk due to a decrease in the capacity proportions (e.g., conditions at presence of measures at *P*_0_ = 10^−3^: 0.056 at capacity proportion of 75%; 0.027 at capacity proportion of 50%; 0.012 at capacity proportion of 25%). Under the presence of preventive measures, the relative risk of infection was almost similar among the three conditions: *P*_0_ = 10^−3^ and the capacity proportion = 25% (relative risk of infection: 0.012); *P*_0_ = 2×10^−4^ and the capacity proportion = 75% (relative risk of infection: 0.011); and *P*_0_ = 10^−4^ and the capacity proportion = 100% (relative risk of infection: 0.0093). Similar results were confirmed for the baseball game condition (Figure S4).

**Figure 2.**
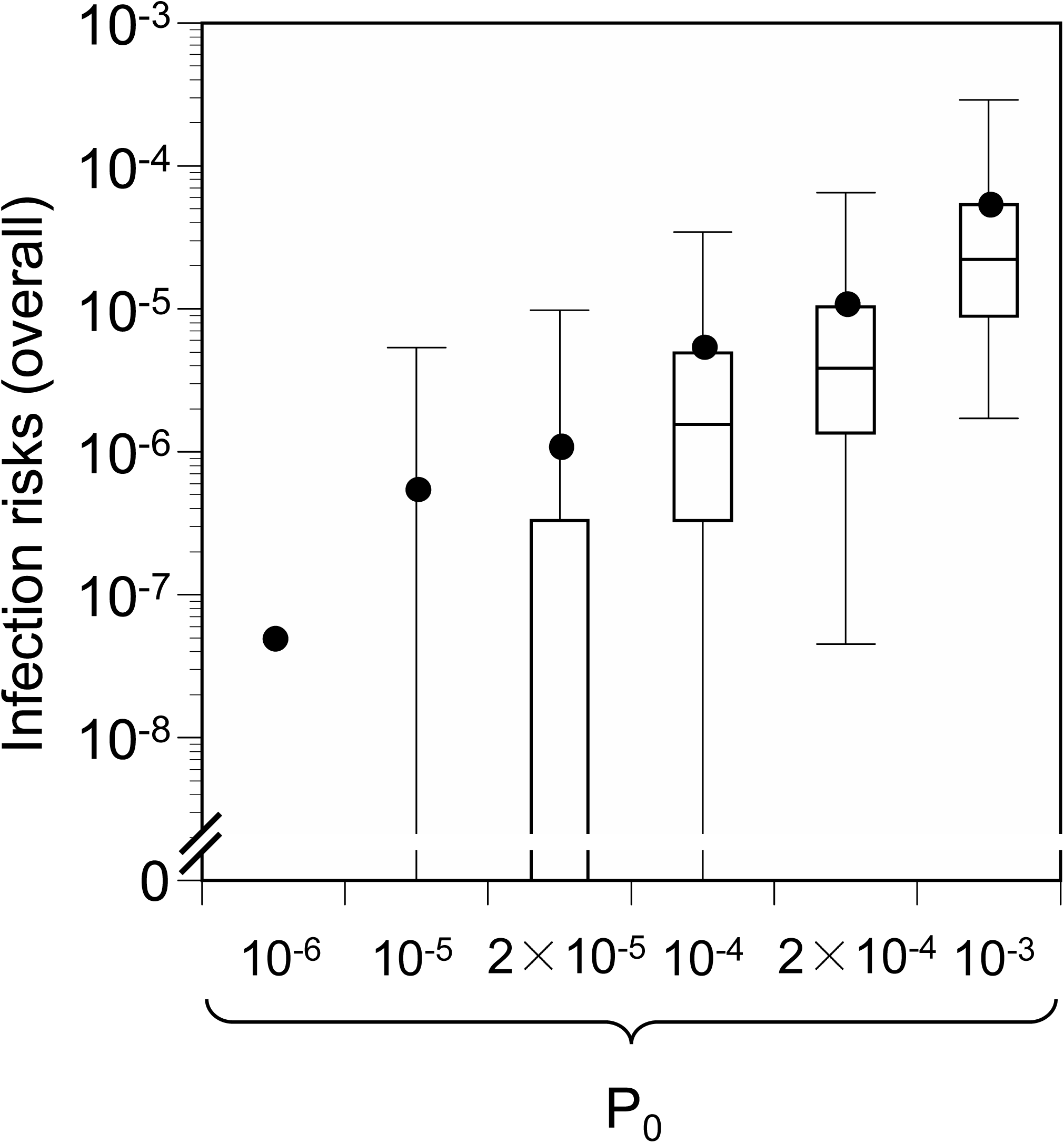
Comparison of infection risk among different crude probabilities of a spectator being an infector (*P*_0_). Stadium capacity = 40,000 persons, capacity proportion 50%, presence of preventive measures (Condition B), football condition. Box-and-whisker plots represent the 2.5, 25, 50, 75, and 97.5 percentiles. Closed circles represent the arithmetic mean of the simulation.

**Figure 3.**
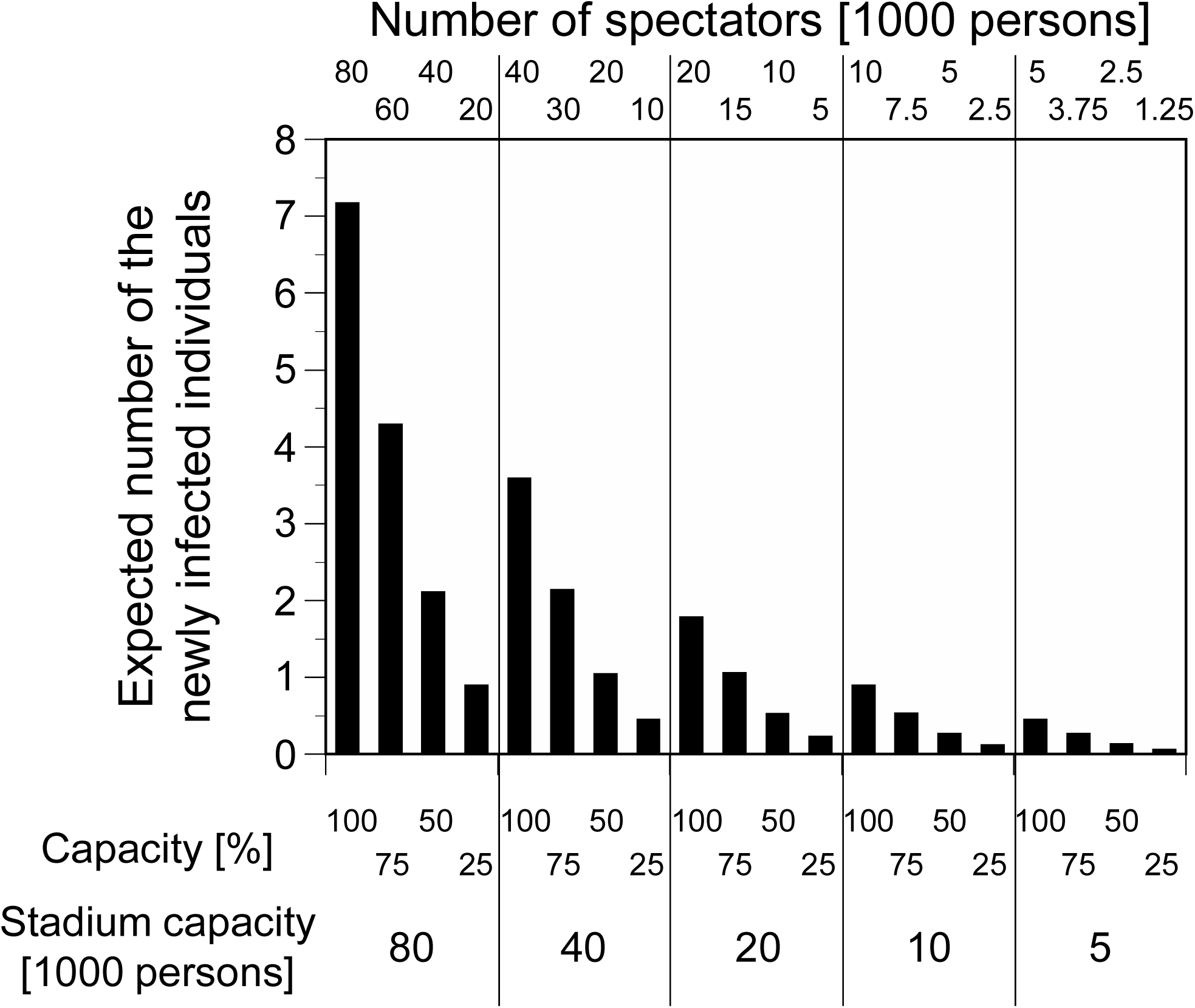
The expected number of newly infected individuals at different stadium capacities and capacity proportions. Presence of preventive measures (Condition B), a crude probability of a spectator being an infector (*P*_0_) = 10^−3^, football condition.

**Figure 4.**
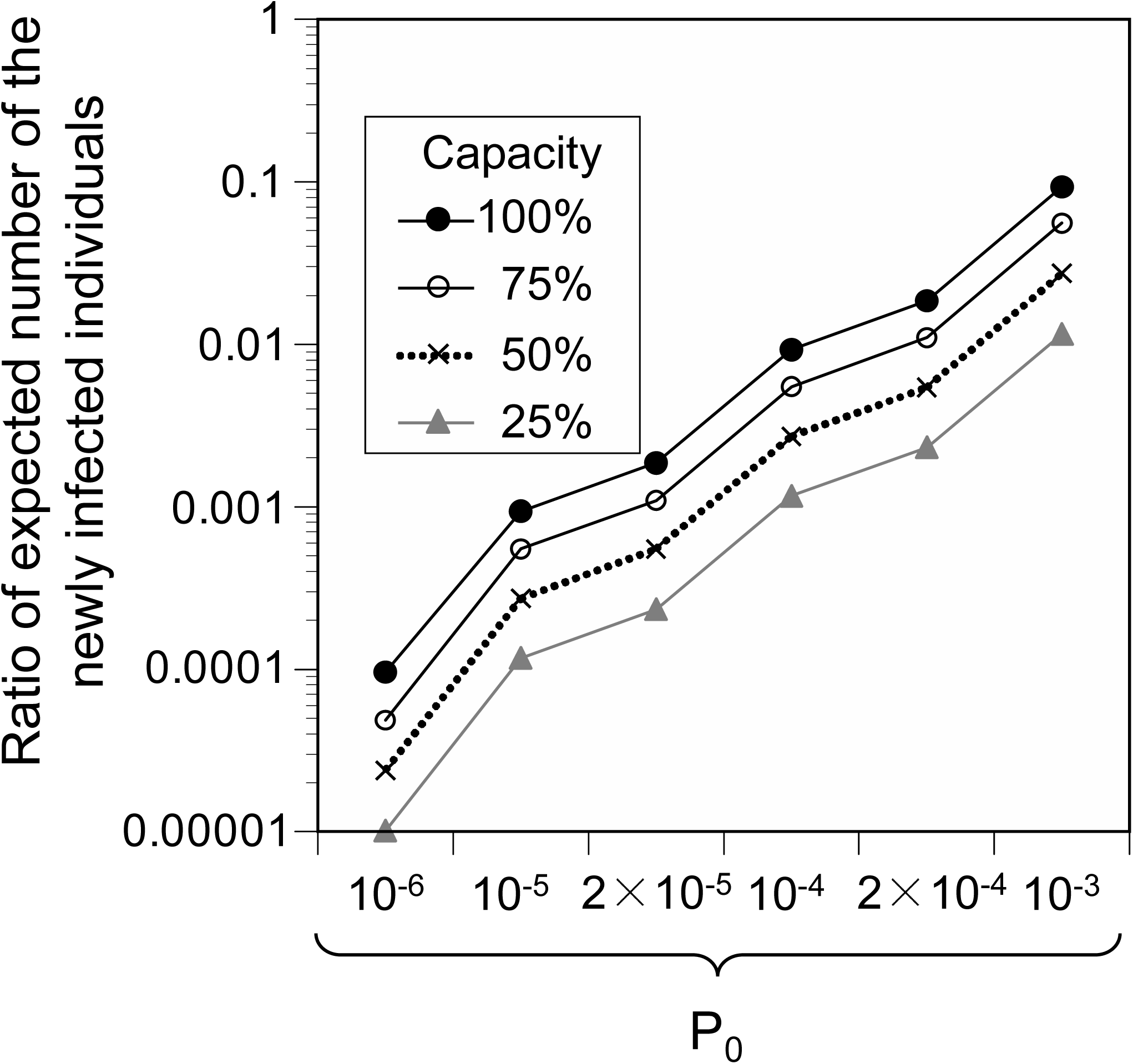
Relative risk of infection with the preventions at different *P*_0_ and capacity proportions compared to the risk of infection at *P*_0_=10^−3^, 100% capacity proportion, and no preventions. Condition B, stadium capacity = 80,000 persons, football condition. *P*_0_: a crude probability of a spectator being an infector.

### 3.3. Estimation of infection risk using *P*_0_, number of spectators, and capacity proportion as explanatory variables

We estimated the partial regression coefficients in the multiple regression analysis with *P*_0_, number of spectators, and capacity proportion as explanatory variables, and the expected number of newly infected individuals as the objective variable under Condition B with the presence of preventive measures (Tables 2 and S2). The regression showed that deviance explained 0.9998 (*P* < 0.001) for both the football game and the baseball game conditions. The expected number of newly infected individuals was significantly associated with *P*_0_, number of spectators, and capacity proportion. The partial regression coefficients calculated by standardizing the explanatory variables were higher for *P*_0_, followed by the number of spectators, and capacity proportion, for both the football and baseball conditions.

The regression equations obtained here and *P*_0_, number of spectators, and the capacity proportion of the actual games were used to estimate the expected number of newly infected individuals for each game (Figure S5). Cumulatively over the periods (from August, 2020 to January 4, 2021 for football games and to November 25, 2020 for baseball games), the number of asymptomatic infectors entering the stadium and the number of newly infected individuals was estimated to be 151.9 persons (0.005%) and 6.4 persons (0.0002%), respectively, among 2,935,947 spectators at football games, and 181.0 persons (0.004%) and 9.5 persons (0.0002%), respectively, among 4,439,258 spectators at baseball games. For football games, the arithmetic mean and maximum expected number of newly infected individuals per game were 0.0073 persons and 0.42 persons, respectively. For baseball games, arithmetic mean and maximum were 0.019 persons and 0.26 persons, respectively. No games exceeded the expected number of newly infected individuals of 1. The percentage of the estimated number of newly infected individuals per game of 0.1 or less was 99.2% for football games and 97.9% for baseball games. Both the cumulative estimated number of infectors entering the stadiums and that of newly infected individuals were higher than the reported numbers (i.e., 5 persons and 0 persons, respectively, for the football games (*8*), and 5 persons and 0 persons, respectively, for the baseball games (*7*)).

**Table 2.** Partial regression coefficients for the infection risk with the presence of preventive measures. Condition B, football condition. Objective variable is the expected number of newly infected individuals. CI: confidence interval.

|  | Partial regression coefficient<br>B (95% CI) | Partial regression coefficient<br>B' (95% CI) <sup>a</sup> |
| --- | --- | --- |
| $\log_{10} P_0$ | 2.310 (2.295–2.326) | 2.246 (2.230–2.261) |
| $\log_{10}$ *Number of spectators* | 2.312 (2.299–2.326) | 1.119 (1.113–1.126) |
| *Capacity proportion* | 0.983 (0.965–1.002) | 0.276 (0.271–0.281) |

<sup>a</sup>Partial regression coefficients were estimated from z-score standardized explanatory variables.

## 4. Discussion

In this study, we evaluated the effect of preventive measures, including face masks, *P*_0_, stadium capacity, and capacity proportion on the infection risk. We further estimated the expected number of newly infected individuals under actual game conditions. At *P*_0_ of 10^−6^–10^−3^, stadium capacity of 5000–80,000 persons, and capacity proportion in the range of 25% to 100%, the infection risk was reduced by 86%–95% with the implementation of face-mask wearing and hand washing measures. Among the individual measures, wearing a face mask was particularly effective, and the infection risk increased as the face-mask-wearing proportion decreased. A face-mask-wearing proportion of 90%, which corresponded to actual game conditions, increased the infection risk by 2.2 times compared to the proportion of 100%. The effectiveness of single measures, excluding face masks, was limited, but this does not mean that each measure is not significant, as found in the high reduction in infection risk due to the combination of all the preventive measures. Other preventive measures effectively reduced the risk that residually remained, even after face-wearing masks.

Regarding the relationship between infection risk and *P*_0_, the infection risk was 1100 times higher when *P*_0_ was 1000 times higher, confirming a roughly linear relationship. In Japan, the infection prevalence differs greatly among prefectures by about 100 to 200 times (e.g., confirmed positive cases among the population of 100 thousand people in 7 days on December 17, 2020: Tokushima Prefecture, 0.14; Tokyo, 28.45 (*35*)). Therefore, even if a game were held with the same stadium capacity and capacity proportion, the expected number of newly infected individuals could vary by 100 to 200 times, depending on the location of the game.

We examined the relationship between the infection risk and the stadium capacity or the capacity proportion, and found that the expected number of newly infected individuals increased as the capacity proportion increased when the stadium capacity was constant. Furthermore, the ratio of the expected number of newly infected individuals at 100% to 25% was 7.9–8.0, which was higher than 4. When the number of spectators (i.e., stadium capacity × capacity proportion) was the same, the infection risk was slightly increased with an increase in capacity proportion. These results imply that the risk of infection per spectator increases as the capacity proportion increases. In this study, we modelled the difference in physical distance among spectators in a stand according to the capacity proportion. The results from this study suggested that the proximity of seats by spectator seating could be one factor in the increased risk of infection. The multiple regression analysis also showed that the expected number of newly infected individuals depended not only on *P*_0_ but also on the number of spectators and the capacity proportion, highlighting the importance of considering the number of spectators and the capacity proportion in infection risk management.

It is beneficial to ensure compliance and effectiveness of preventive measures at mass gathering events because the reduction effect of measures is remarkable. When mitigating the number of spectators and the capacity proportion, it is expected that the event will be implemented in accordance with a local infection prevalence. For example, this study showed similar infection risk levels with a stadium capacity of 80,000 persons among a capacity proportion of 25% at *P*_0_ = 10^−3^, 75% at 2×10^−4^, and 100% at 10^−4^. Setting the capacity proportion according to the local infection prevalence allowed us to accept the event with the same risk level of newly infected individuals. Considering the fact that spectators’ viewing patterns differ depending on the type of mass gathering event, an assessment based on an actual condition and its correspondence is required.

The expected number of infectors entering the stadiums and the expected number of newly infected individuals estimated in this study were larger than the values actually reported. A possible reason for the former is that the identification of infectors entering the stadium for the games relies on voluntary reports from infectors. Therefore, the actual number of infectors entering the stadium may have not been fully captured. Another possibility is that the infection prevalence among the spectators entering the stadium was lower than that among the entire population, owing to differences in age structure and health attributes.

It should be noted that the results of the expected number of newly infected individuals simulated in this study might be underestimated due to the model settings regarding the face-mask-wearing proportion and *P*_0_. Nevertheless, we found a gap in the expected number of newly infected individuals between the estimation (6.4 persons in the football games and 9.5 persons in the baseball games) and the actual reports (0 for both games). There are four possible reasons for this observation. First, the number of infectors who actually entered the stadium was low, for the reasons mentioned earlier. Second, the risk assessed by the model in this study showed overestimation. For example, the actual risk might be lower than the infection risk assessed by the model, because vocal cheering has been prohibited in professional football and baseball games. Third, the estimated and reported values of the number of newly infected individuals were less than approximately two millionths of the number of spectators; therefore, the infection risk level was too small to accurately estimate under the presence of some uncertainties in the model. Fourth, as with the number of infectors entering stadiums described above, it is possible we missed capturing the actual newly infected individuals. Limited testing and the presence of asymptomatic individuals might also contribute to this miss.

Although empirical epidemiological studies of the number of infected individuals at mass gathering events are underway (*9, 10*), epidemiological estimates of infection risk are difficult to make owing to the small number of newly infected individuals. Further accumulation of empirical cases and evaluations in actual mass-gathering events is necessary to refine the infection risk assessment. In this study, despite the aforementioned uncertainties, the combination of the model and data from actual games showed that there were few new infected individuals among approximately 3–4 million spectators. The findings of the study will be useful in decision-making regarding measures to be taken for events during infectious disease pandemics.

## Data Availability

All data can be found in the text.

## Acknowledgement

We thank Dr. Fuminari Miura (Center for Marine Environmental Studies, Ehime University) for his help for the model development and implementation.

## Funds

No external financial support is used for this article.

---

Table S1. Reduction ratio of infection risks due to preventions at different *P*_0_, stadium capacities, and capacity proportions. Condition B, baseball condition. *P*_0_: a crude probability of a spectator being an infector.

Table S2. Partial regression coefficients for infection risk with the presence of preventive measures. Condition B, baseball condition. Objective variable is the expected number of newly infected individuals. CI: confidence interval.

Figure S1. Sensitivity analysis for changes in virus concentrations in saliva. (a) Absence of preventions; (b) presence of preventions. Condition B, a crude probability of a spectator being an infector (*P*_0_) =10^−3^, stadium capacity = 40,000 persons, capacity proportion = 50%, football condition. “×1” represents the reference scenario. “×2” represents a viral load that increased to 2 times the reference scenario. Box-and-whisker plots represent the 2.5, 25, 50, 75, and 97.5 percentiles. Closed circles represent the arithmetic mean of the simulation.

Figure S2. Comparison of infection risk among different crude probabilities of a spectator being an infector (*P*_0_). Stadium capacity = 40,000 persons, capacity proportion 50%, presence of preventions (Condition B), baseball condition. Box-and-whisker plots represent the 2.5, 25, 50, 75, and 97.5 percentiles. Closed circles represent the arithmetic mean of the simulation.

Figure S3. The expected number of newly infected individuals at different stadium capacities and capacity proportions. Presence of preventions (Condition B), a crude probability of a spectator being an infector (*P*_0_) = 10^−3^, baseball condition.

Figure S4. Relative risk of infection with the preventions at different *P*_0_ and capacity proportions compared to the risk of infection at *P*_0_=10^−3^, 100% capacity proportion, and no preventions. Condition B, stadium capacity = 80,000 persons, baseball condition. *P*_0_: a crude probability of a spectator being an infector.

Figure S5. The expected number of newly infected individuals, estimated from regression analysis. Presence of preventions (Condition B). No spectator games were excluded. (a) The relationship between the expected number of newly infected individuals and the number of confirmed cases at sites and the number of spectators (football condition); (b) the relationship between the expected number of newly infected individuals and the number of confirmed cases at sites and the number of spectators (baseball condition); (c) frequency of the expected number of newly infected individuals (football condition); (d) frequency of the expected number of newly infected individuals (baseball condition). The legends in Figures (a) and (b) represent the expected number of newly infected individuals.

