## supplemental for "Assessment of COVID-19 risk and prevention effectiveness among spectators of mass gathering events"

Table S1.

| Stadium capacity | Capacity proportion | Number of spectators | *P*_0_ | | | | | |
| --- | --- | --- | --- | --- | --- | --- | --- | --- |
|  |  |  | 10^-6^ | 10^-5^ | 2×10^-5^ | 10^-4^ | 2×10^-4^ | 10^-3^ |
| 80000 | 100% | 80000 | 90% | 90% | 90% | 90% | 90% | 90% |
|  | 75% | 60000 | 92% | 91% | 91% | 91% | 91% | 91% |
|  | 50% | 40000 | 93% | 93% | 93% | 93% | 93% | 93% |
|  | 25% | 20000 | 94% | 93% | 93% | 93% | 93% | 93% |
| 40000 | 100% | 40000 | 90% | 90% | 90% | 90% | 90% | 90% |
|  | 75% | 30000 | 92% | 91% | 91% | 91% | 91% | 91% |
|  | 50% | 20000 | 94% | 93% | 93% | 93% | 93% | 93% |
|  | 25% | 10000 | 93% | 93% | 93% | 93% | 93% | 93% |
| 20000 | 100% | 20000 | 91% | 90% | 90% | 90% | 90% | 90% |
|  | 75% | 15000 | 92% | 91% | 91% | 91% | 91% | 91% |
|  | 50% | 10000 | 93% | 93% | 93% | 93% | 93% | 93% |
|  | 25% | 5000 | 95% | 94% | 93% | 93% | 93% | 93% |
| 10000 | 100% | 10000 | 90% | 90% | 90% | 90% | 90% | 90% |
|  | 75% | 7500 | 91% | 92% | 91% | 91% | 91% | 91% |
|  | 50% | 5000 | 94% | 93% | 93% | 93% | 93% | 93% |
|  | 25% | 2500 | 94% | 94% | 94% | 93% | 93% | 93% |
| 5000 | 100% | 5000 | 91% | 91% | 90% | 90% | 90% | 90% |
|  | 75% | 3750 | 91% | 91% | 92% | 91% | 91% | 91% |
|  | 50% | 2500 | 93% | 93% | 93% | 93% | 93% | 93% |
|  | 25% | 1250 | 94% | 94% | 94% | 93% | 93% | 93% |

Table S2.

|  | Partial regression coefficient B (95% CI) | Partial regression coefficient B’ (95% CI) ^a^ |
| --- | --- | --- |
| log_10_ *P*_0_ | 2.322 (2.309 – 2.336) | 2.257 (2.244–2.270) |
| log_10_ *Number of spectators* | 2.314 (2.302 – 2.325) | 1.120 (1.114–1.125) |
| *Capacity proportion* | 1.071 (1.055 – 1.086) | 0.300 (0.296–0.305) |

^a^ Partial regression coefficients were estimated from z-score standardized explanatory variables.

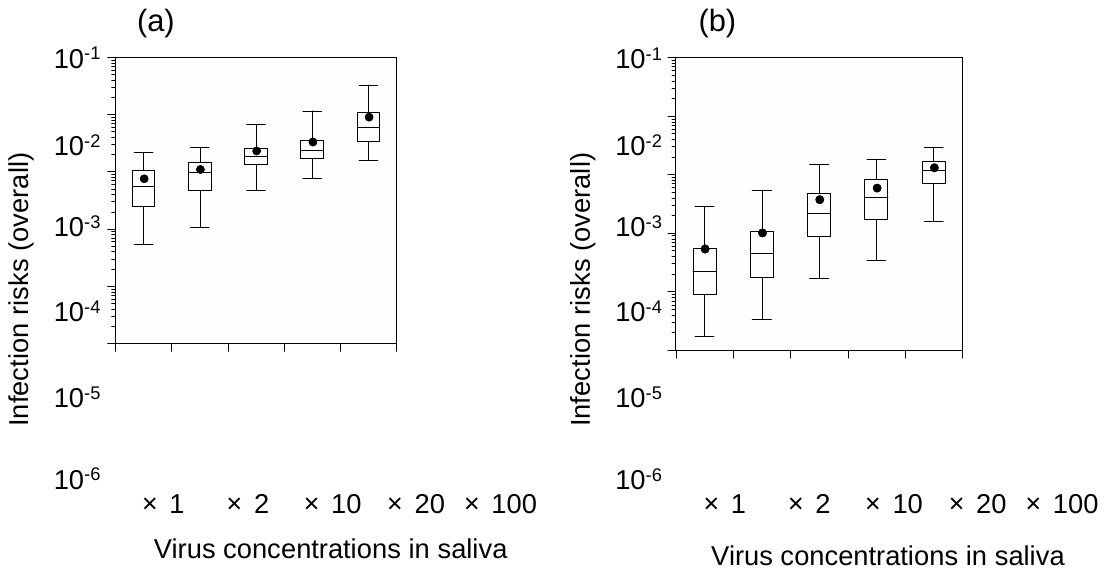

Figure S1.

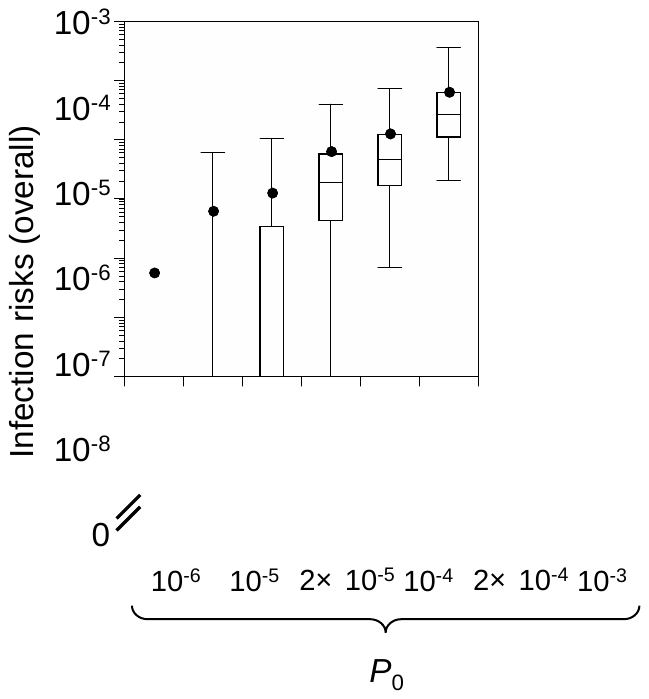

Figure S2.

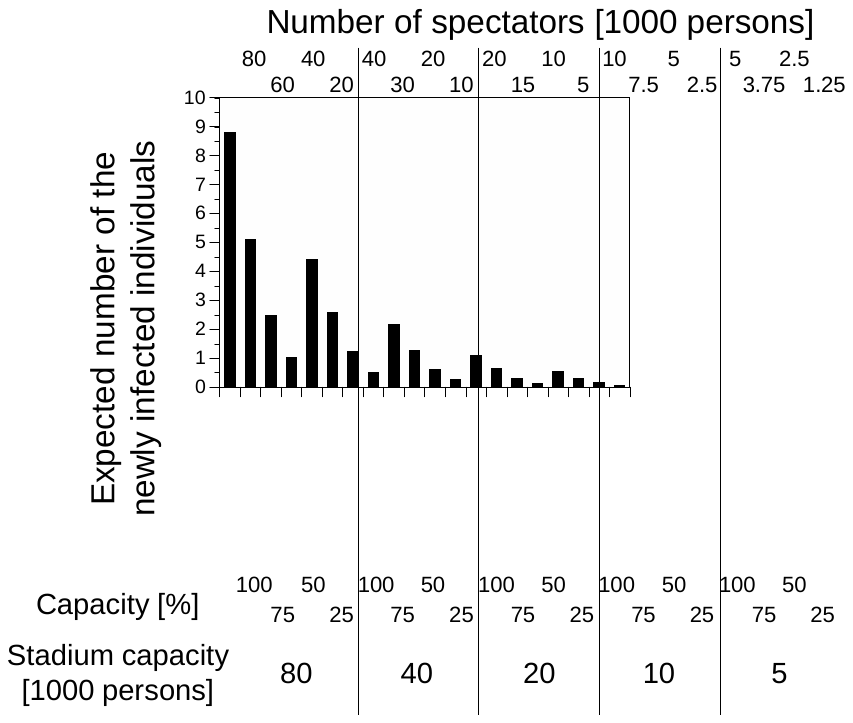

Figure S3.

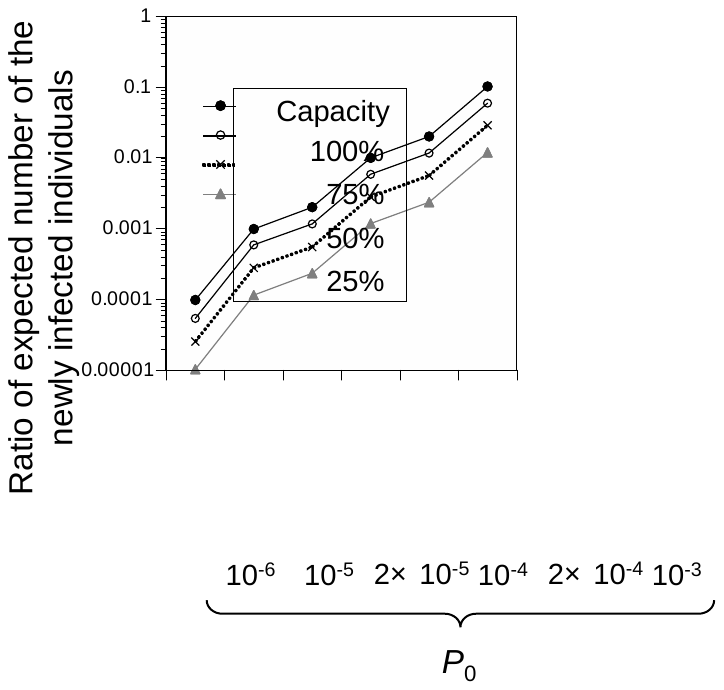

Figure S4.

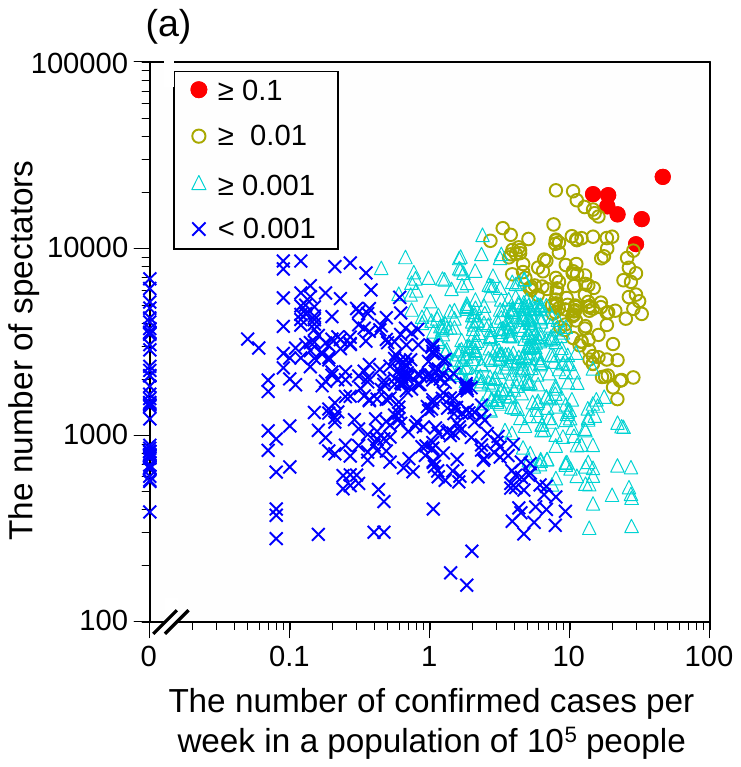

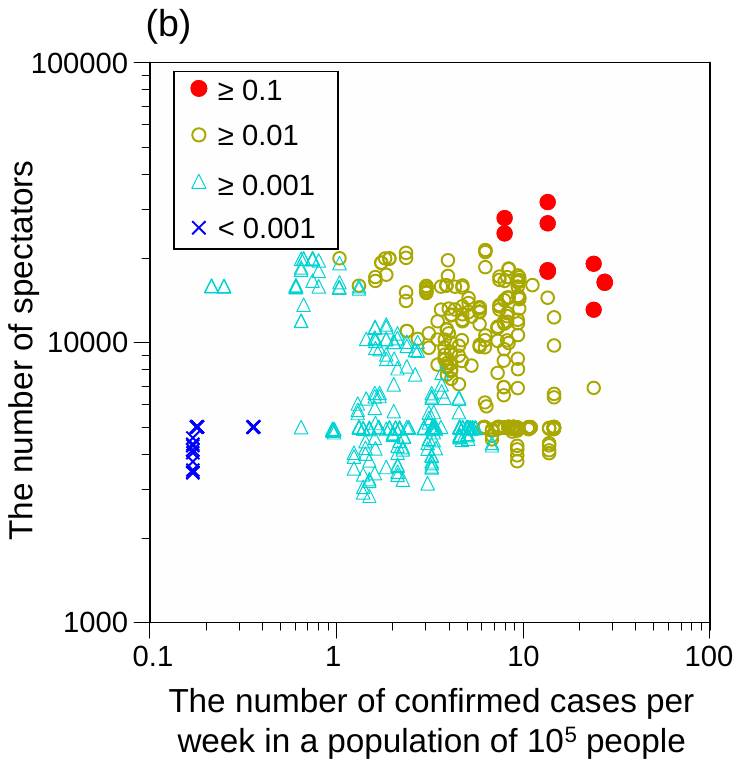

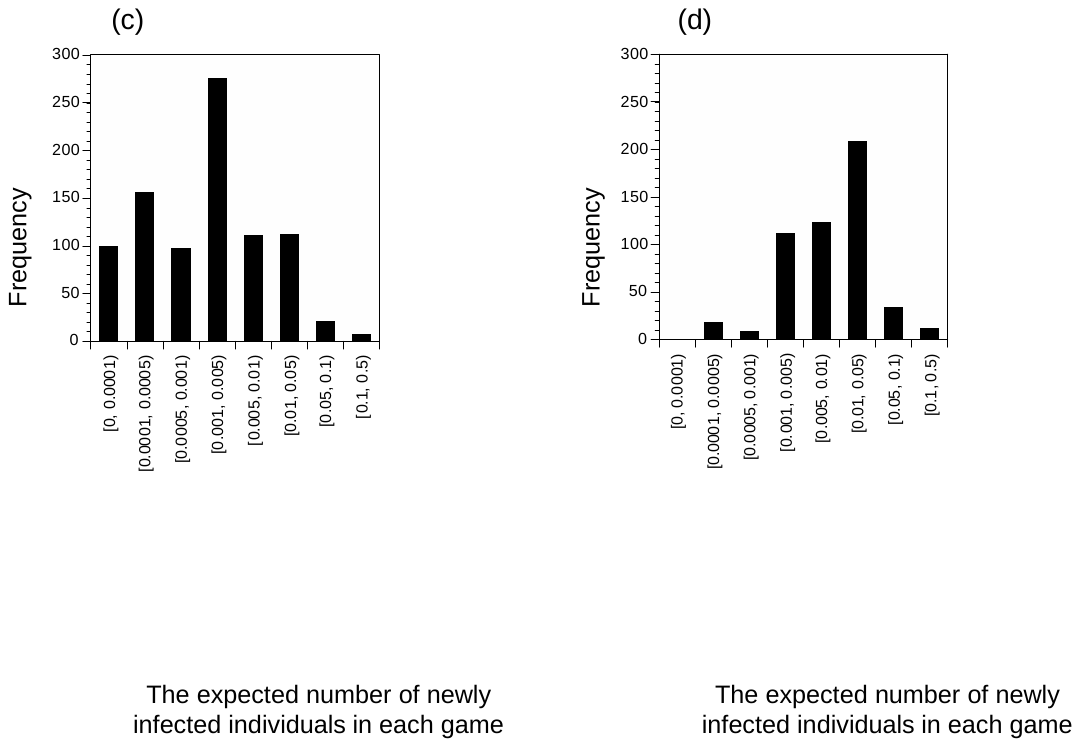

Figure S5.
